# Deep learning enhanced magnetocardiography enables multi-task detection of coronary, ventricular, and rhythm disorders

**DOI:** 10.64898/2025.11.30.25341301

**Authors:** Dominik D. Kranz, Oruç Kahriman, Dominic Dischl, Sascha Treskatsch, André Sander, Johannes Brachmann, Jai-Wun Park, Niels Wessel

**Affiliations:** Berlin Institute of Health at Charité – Universitätsmedizin Berlin, Germany; Charité Universitätsmedizin Berlin, Germany; Department of Physics, Humboldt-Universität zu Berlin, Germany; ID Information und Dokumentation im Gesundheitswesen GmbH & Co. KGaA; German Heart Centre Munich, Germany; Clinic for Anaesthesiology and Intensive Care, Campus Benjamin Franklin, Charité Universitätsmedizin Berlin, Germany; Sana Medical School, Coburg, Germany; University of Split School of Medicine; German Heart Center, DHZC at Charité – Universitätsmedizin Berlin, Germany; MSB Medical School Berlin, Germany

## Abstract

**Background:** Magnetocardiography (MCG) captures the magnetic fields generated by myocardial currents, theoretically preserving electrophysiological details lost in surface potentials. However, its clinical application has been hindered by small datasets and the complexity of analysing high-dimensional magnetic field data. We sought to develop a self-supervised deep learning framework to detect high burden coronary artery disease (CAD), left ventricular dysfunction, and arrhythmia risk from resting MCG recordings.

**Methods:** We developed MCG2Vec, a contrastive deep learning encoder trained on raw 64-channel MCG signals. The model was pre-trained on unlabeled 10-second segments to learn generalizable signal morphology, then fine-tuned for clinical tasks in a retrospective cohort of 1,732 consecutive patients. The primary endpoints were the detection of significant CAD (_≥_ 70% stenosis), reduced left ventricular ejection fraction (LVEF <55%), and atrial fibrillation (AF) risk, all derived from sinus-rhythm recordings. Model performance was evaluated using patient-stratified five-fold cross-validation and interpreted using Grad-CAM activation mapping.

**Results:** In the validation analysis, the model detected significant CAD with an area under the curve (AUC) of 0.89 (95% CI 0.82 - 0.93) and successfully localized ischemia to the left anterior descending (AUC 0.88), right coronary (0.82), and left circumflex arteries (0.82). Reduced LVEF was identified with an AUC of 0.81 (95% CI 0.71 - 0.91). Furthermore, the model predicted a history of paroxysmal AF from sinus-rhythm recordings with an AUC of 0.77 (95% CI 0.69 - 0.85). Explainability analysis confirmed that the model relied on physiologically distinct phases of the cardiac cycle (repolarization heterogeneity for ischemia and P-wave/S-wave morphology for arrhythmia risk) rather than non-specific noise.

**Conclusion:** Deep learning-enhanced magnetocardiography enables the accurate, non-invasive detection of ischemia, ventricular dysfunction, and arrhythmia risk from a single resting scan. By unlocking the latent diagnostic information within the cardiac magnetic field, this approach offers a scalable, radiation-free adjunct to standard electrocardiography for precision cardiac diagnostics.

## Introduction

Cardiovascular disease (CVD) remains the leading cause of morbidity and mortality worldwide, underscoring the need for accurate, accessible, and non-invasive diagnostic tools capable of identifying disease at an early, potentially reversible stage [1], [2], [3]. Despite major technological advances, current cardiovascular diagnostics still rely on a trade-off between simplicity and precision [4]. At one end, electrocardiography (ECG) remains the global first-line tool due to its ubiquity and low cost [4]. However, the ECG records surface electrical potentials rather than the true three-dimensional myocardial electrical field [5], limiting sensitivity and specificity for ischemia, arrhythmogenic substrates, and subtle myocardial dysfunction [4], [6]. The exercise ECG, once central to ischemia assessment, has also lost relevance due to poor accuracy in multivessel disease, microvascular dysfunction, and atypical symptom profiles [4].

At the opposite end of the diagnostic spectrum, techniques such as coronary angiography, CT angiography, and nuclear perfusion imaging deliver detailed anatomical or perfusion information but involve radiation exposure, contrast agents, procedural risks, and limited accessibility [4], [7]. Even cardiac MRI, while non-invasive and radiation-free, remains resource-intensive and typically limited to tertiary centers [8]. This leaves a diagnostic gap between inexpensive but coarse tools and high-fidelity, invasive, or resource-heavy modalities. No scalable, non-invasive modality currently offers high-fidelity electrophysiological information without radiation, contrast, or operator dependence [9], [10].

From a biophysical standpoint, ECG limitations stem from its reliance on conductive tissue pathways: electrical potentials are attenuated and distorted as they traverse skin, muscle, and bone [5], [9]. MCG in contrast, measures the magnetic fields generated by cardiac electrical currents, which propagate through tissue with minimal distortion and therefore preserve electrophysiological detail [9], [10], [11].

Recent advances in self-supervised and contrastive learning provide a path to unlock the high-dimensional MCG signal space by learning generalizable representations directly from raw recordings [12], [13], [14]. These approaches offer the potential to extract subtle, physiologically relevant information that conventional analysis has been unable to exploit [15], [16].

Here, we develop an AI-enabled MCG framework that combines the biophysical fidelity of MCG with contrastive representation learning. We train a self-supervised encoder on raw 64-channel MCG recordings and fine-tune it for clinically meaningful tasks: identifying ischemia in patients with suspected high burden chronic coronary artery disease (CAD), estimating left ventricular ejection fraction (LVEF) as a marker of systolic dysfunction, and detecting atrial fibrillation (AF) risk from sinus-rhythm recordings. We further evaluate spatial learning by predicting stenoses of the left anterior descending (LAD), right coronary artery (RCA), and left circumflex artery (LCX) based on sensor-array positioning.

These tasks reflect high-stakes, real-world diagnostic challenges. Coronary artery disease remains the leading global cause of morbidity and mortality [1], and current non-invasive tools lack adequate accuracy for many patients who ultimately undergo angiography [4]. Heart failure affects millions worldwide, and timely recognition of reduced LVEF remains essential yet operator-dependent with conventional imaging [17], [18]. AF is common, carries major thromboembolic and heart-failure risk, and often remains undetected or intermittent [19]. By targeting these scenarios, our work positions MCG not as a replacement for ECG, but as a scalable, radiation-free adjunct that has the potential to improve non-invasive risk stratification, reduce unnecessary invasive procedures, and enable earlier intervention. Our results are consistent with the renewed momentum in magnetocardiography research, reflected in recent diagnostic advances and editorials calling for a re-evaluation of MCG’s clinical potential [20], [21], [22], [23].

## Methods

### Study design and population

A total of 1,732 consecutive patients underwent clinical MCG examination at the Cardiology Department of Asklepios Hospital Hamburg, Germany. MCG was performed as part of the standard diagnostic work-up for suspected cardiovascular disease. The study was approved by the local ethics committee (Ärztekammer Hamburg, MC-017/09).

Patients were included if (i) an MCG recording of sufficient technical quality was available and (ii) a corresponding clinical reference test existed for at least one of the three diagnostic tasks. Exclusion criteria included acute coronary syndrome, incomplete clinical documentation, missing reference standards, or insufficient MCG signal quality.

Three task-specific cohorts were defined:

1. CAD label construction In the source clinical dataset, stenosis severity was recorded only as binary per-vessel indicators for the LAD, RCA, and LCX (0 = <70%, 1 = _≥_70%). Because this encoding does not distinguish between true absence of stenosis and moderate stenosis (e.g., 30–60%), a simple “any-vessel” definition of CAD would have grouped patients with mild or intermediate disease into the negative class. To reduce label noise, we therefore defined high-burden obstructive coronary artery disease (CAD) as the presence of _≥_70% stenosis in at least two major epicardial vessels. Patients were assigned to the negative class if all three vessel indicators were 0 (i.e., no vessel reached the _≥_70% threshold), acknowledging that this group may still include intermediate non-obstructive disease below the binary cutoff. Vessel-level labels (LAD, RCA, LCX _≥_70%) were derived directly from the corresponding binary indicators for the localization task.
2. LVEF Patients with a transthoracic echocardiogram performed within the clinically accepted temporal window were included in the LVEF cohort. Reduced systolic function was defined as LVEF < 55% in accordance with ESC heart failure guidelines. Patients with uninterpretable echocardiograms or missing LVEF measurements were excluded.
3. AF AF status was determined from structured clinical documentation and MCG records. Only sinus-rhythm MCG recordings were used. Patients were labelled as “known AF” if they had documented paroxysmal AF at any point in their medical history, and “no known AF” otherwise. Individuals with uncertain or ambiguous clinical AF history were excluded. All recordings were manually checked by a medical professional to be in sinus rhythm, and AF rhythms were excluded.

The timing between MCG and reference tests reflected real-world clinical practice. Patient flow for all three tasks, including exclusions and cohort sizes, is summarized in Figure 1.

**Figure 1.**
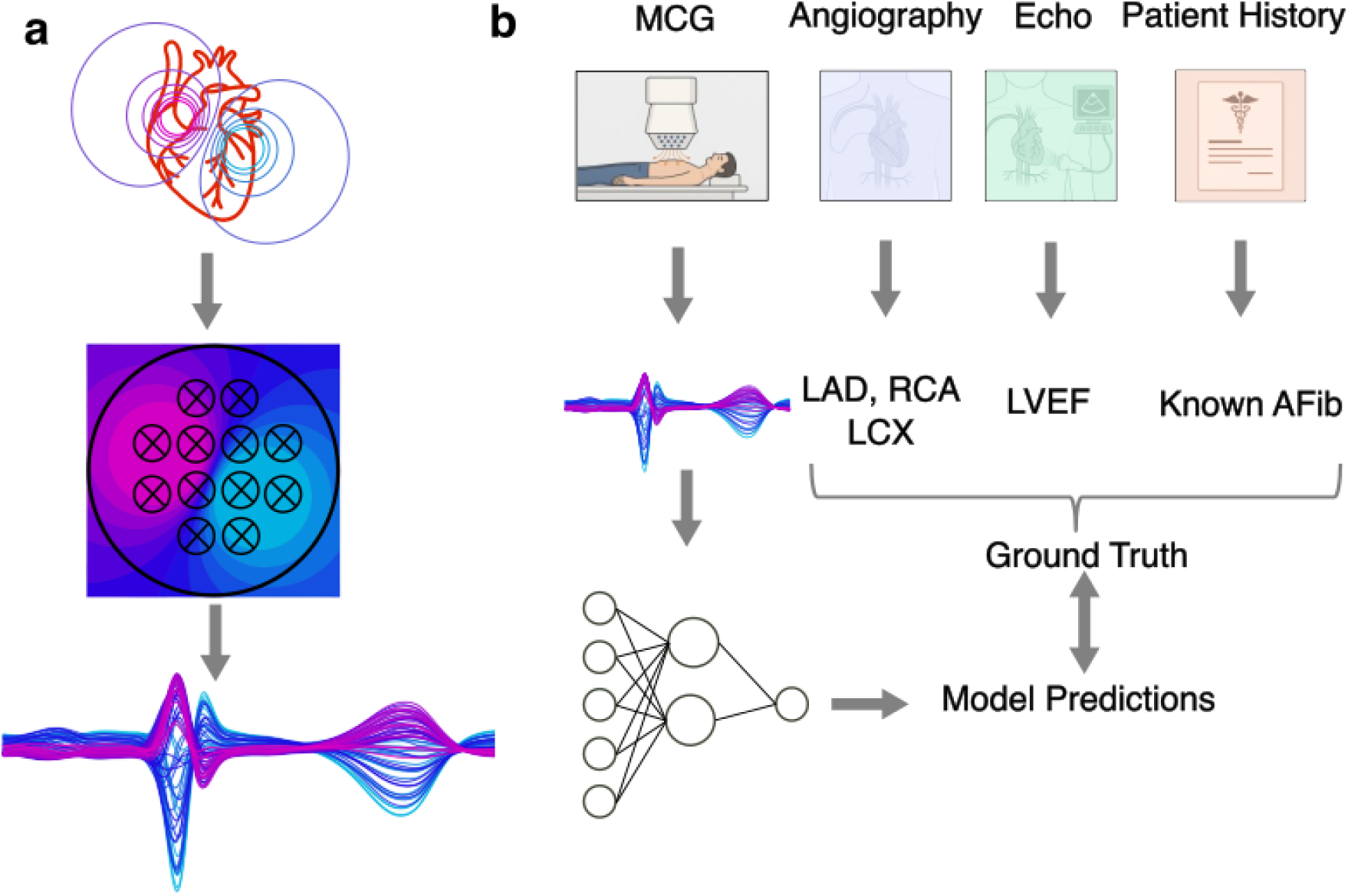
Method visualisation. **(a)** Biophysical principle of MCG. Cardiac electrical activity generates magnetic fields that are captured by a 64-channel SQUID sensor array positioned above the anterior thorax, yielding multichannel MCG waveforms. **(b)** Overview of the supervised learning tasks. MCG recordings were paired with clinical reference standards: coronary angiography for vessel-level stenoses (LAD, RCA, LCX), echocardiography for LVEF, and patient records/ECG documentation for known AF. These labels served as ground truth for model training and evaluation.

### MCG Acquisition and Preprocessing

Magnetocardiography (MCG) recordings were obtained using a 64-channel, second-generation SQUID system (CS-MAG II, Biomagnetik Park GmbH, Germany). Sensors were arranged in a circular grid positioned above the anterior thorax with the patient lying supine. The system was operated in a magnetically shielded environment and recorded cardiac magnetic fields at a sampling rate of 500 Hz using hardware bandpass filtering as provided by the manufacturer. Recording length ranged from 1–4 minutes per patient.

MCG recordings were segmented into non-overlapping 10-second windows at the native sampling rate of 500 Hz. To preserve the 64-channel sensor geometry, channels missing due to technical issues were replaced with zero-valued traces at the corresponding positions. Segments containing flat-line channels or incomplete data were discarded.

Signals underwent standardized filtering consisting of baseline drift removal, 50 Hz power-line suppression, and low-pass filtering to reduce high-frequency noise, all applied in zero-phase mode to preserve waveform morphology. For each segment, a signal-to-noise ratio (SNR) was estimated across channels, and segments with mean SNR < 5 were excluded. Accepted segments were stored as fixed-size 64×5000 matrices for downstream model training.

### Neural network model

**Figure 2.**
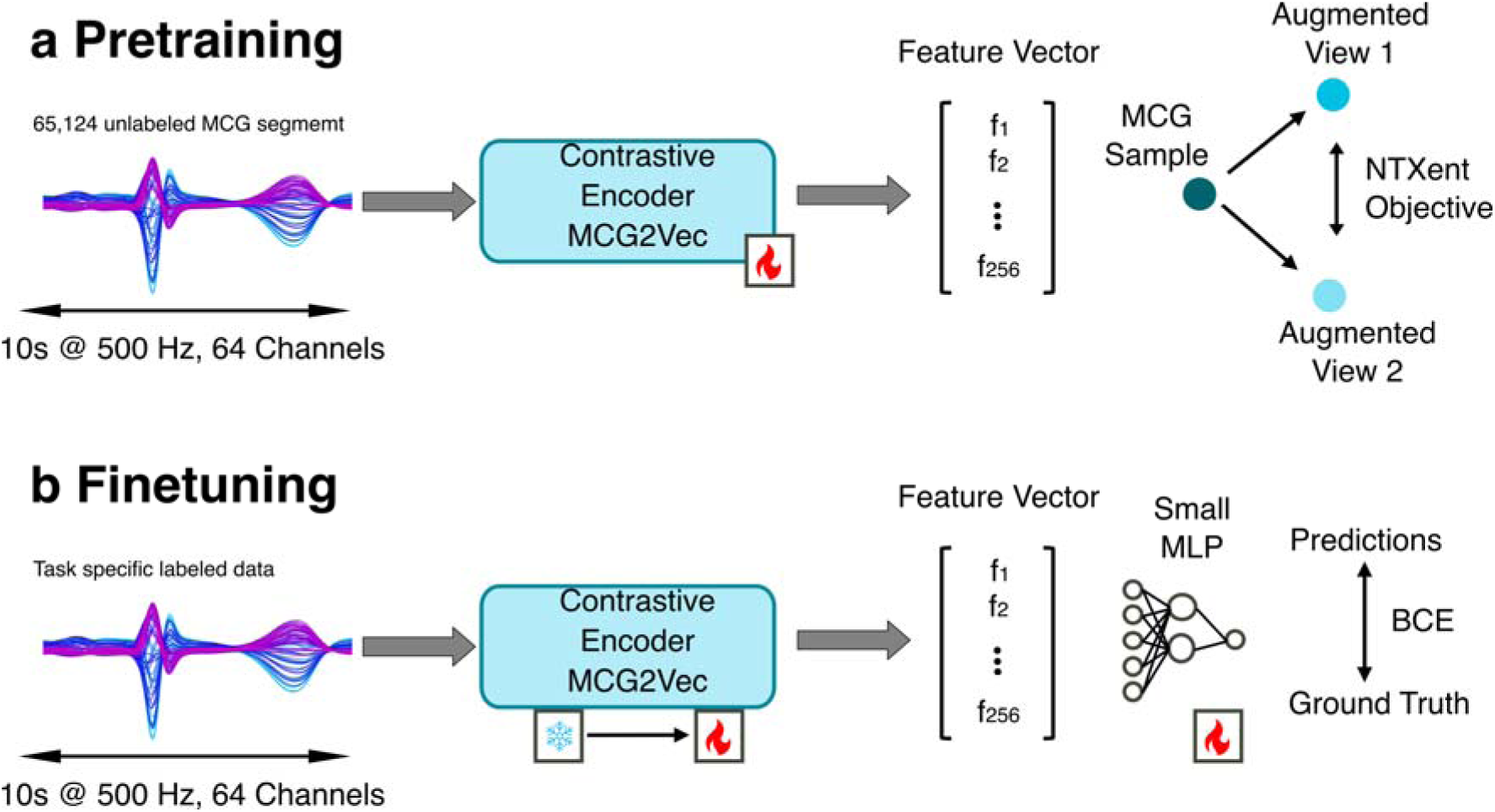
Overview of the MCG encoder model pretraining and finetuning pipeline. During pretraining, short multichannel MCG segments undergo stochastic augmentations and are encoded by a contrastive encoder (MCG2Vec, **a**). The encoder is optimized using a contrastive loss to produce latent embeddings that maximize similarity between augmented views of the same signal while minimizing similarity to embeddings from different signals. In downstream tasks, the pretrained encoder is first frozen then fine-tuned and coupled to task-specific classifiers (**b)**.

### Self-supervised pretraining of the MCG encoder

We first pretrained a convolutional encoder (MCG2Vec) on all available MCG segments using a SimCLR-style contrastive objective. Ten-second MCG windows (64 channels, 5000 samples) were fed to the network as two independently augmented “views”. Augmentations consisted of (i) additive Gaussian noise scaled to the per-channel standard deviation, (ii) random channel dropout (10% of channels replaced by noise drawn from their own mean and variance), and (iii) per-channel temporal masking in which a random contiguous segment of up to ∼75% of the window was replaced by the channel mean.

Pretraining was performed for 100 epochs with a global batch size of 32×number of GPUs using data-parallel training on multiple GPUs. We used the NT-Xent contrastive loss with temperature 0.1, optimized with AdamW (initial learning rate 1×10^-3^, cosine decay over the full training run, final learning rate 1% of the initial value, weight decay 1×10^-4^). The resulting encoder weights were saved and used to initialize all downstream task models.

Contrastive learning was selected because it enforces invariance to channel-specific noise, missing-channel patterns, and non-physiological artifacts—an essential property for MCG, where channel failures are common.

### Downstream fine-tuning for clinical tasks

For each diagnostic task (CAD, LVEF, AF and vessel-level stenosis), we attached a shallow classifier head to the pretrained encoder. Ten-second MCG windows served as inputs (shape 5000×64). The encoder output was passed through dropout 0.2, a 64-unit dense layer with swish activation, and a final sigmoid output neuron.

Fine-tuning was performed within 5-fold patient-level stratified cross-validation. In each fold, all segments from a given patient were assigned exclusively to either the training or validation set. The same data augmentations as in pretraining were applied to training segments only. We used binary cross-entropy loss with snippet-level class weights inversely proportional to class frequency and optimized with Adam. Each fold began with a 5-epoch warm-up phase with the encoder frozen (learning rate 1×10^-3^), followed by joint fine-tuning of encoder and classifier for 30 epochs (learning rate 3×10^-3^).

### Interpretability and Physiological Validation

To ensure that the deep learning model relied on biologically relevant electrophysiological features rather than non-specific noise or acquisition artifacts, we performed a post-hoc interpretability analysis using Gradient-weighted Class Activation Mapping (Grad-CAM). This technique generates a temporal “importance map” for each recording, highlighting the specific segments of the MCG signal that were most influential in determining the model’s diagnosis.

To correlate these machine-learned features with standard cardiac intervals, we aligned the importance maps to the R-peaks of the corresponding cardiac cycles. These beat-aligned maps were then averaged across the validation cohort to project a representative “attention profile” onto a standard template heartbeat. This approach allowed us to visualize whether the model focused on physiologically distinct phases of the cardiac cycle—such as the QRS complex for ventricular depolarization or the ST-segment and T-wave for repolarization—and to verify that the learned features were consistent with known pathophysiological mechanisms of ischemia, systolic dysfunction, and atrial remodeling.

## Results

### Cohort description

From a total of 1,732 consecutive patients undergoing clinical MCG, task-specific cohorts were derived based on the availability of reference standards and signal quality (Figure 3). For the detection of ischemia, the CAD cohort included 208 patients, comprising 87 with high-burden obstructive disease and 121 controls. Patients with significant CAD were older (71.8 ± 10.7 vs. 63.8 ± 15.2 years, p < 0.05) and predominantly male (75% vs. 42%, p < 0.05) compared to those without obstructive disease (Table 1). The LVEF cohort consisted of 238 patients; those with reduced systolic function (n = 95) were similarly older and had a higher proportion of males compared to patients with preserved ejection fraction (n = 143). For the arrhythmia prediction task, 454 patients with sinus-rhythm MCG recordings were analysed. Patients with a documented history of atrial fibrillation (n = 164) were significantly older than controls (71.3 ± 9.8 vs. 65.3 ± 14.3 years) and presented less frequently with acute chest pain (12% vs. 20%).

**Figure 3.**
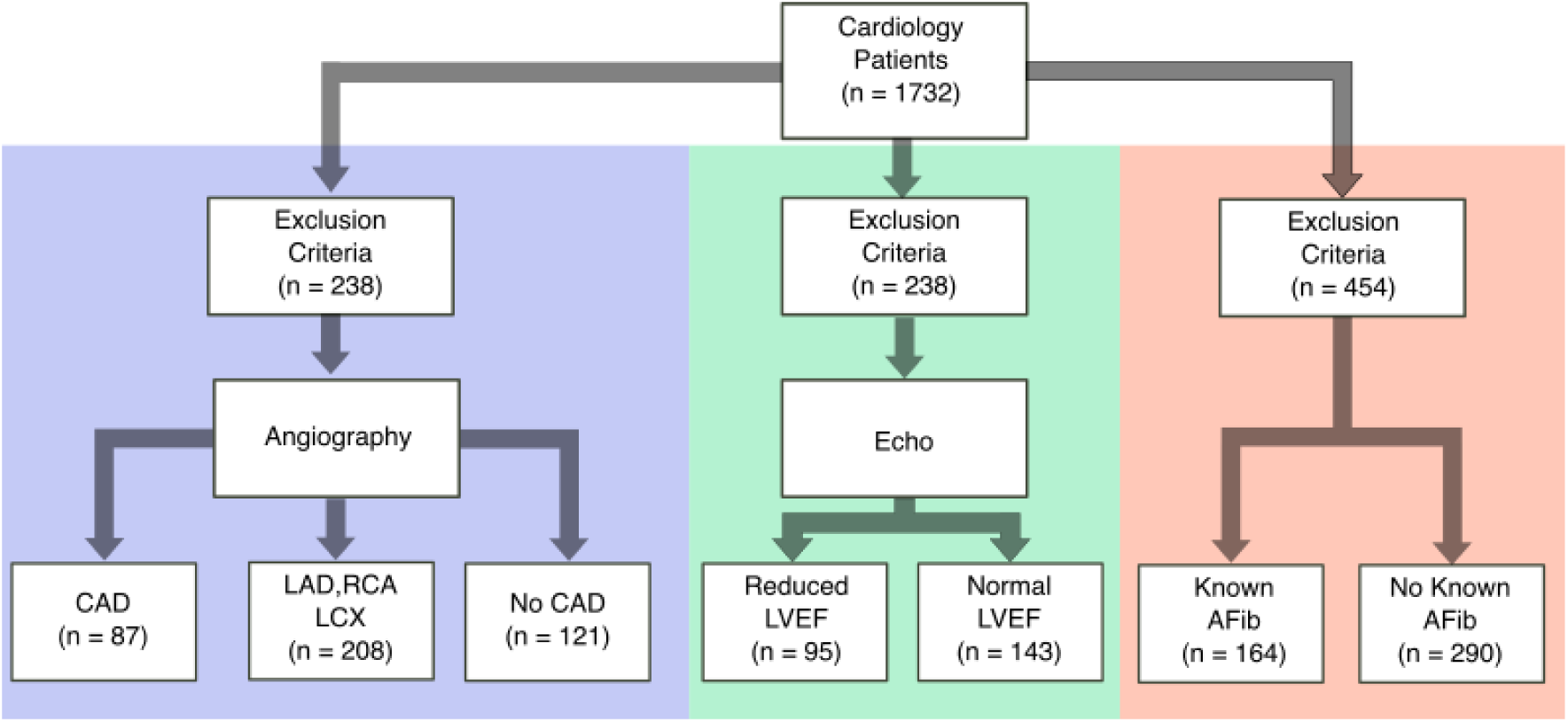
Patient flow for each diagnostic task. Of 1,732 patients with clinical MCG recordings, task-specific cohorts were derived based on availability of corresponding reference tests: CAD (n = 87 with high burden stenosis; n = 208 LAD/RCA/LCX labels; n = 121 no-CAD)

**Table 1.**
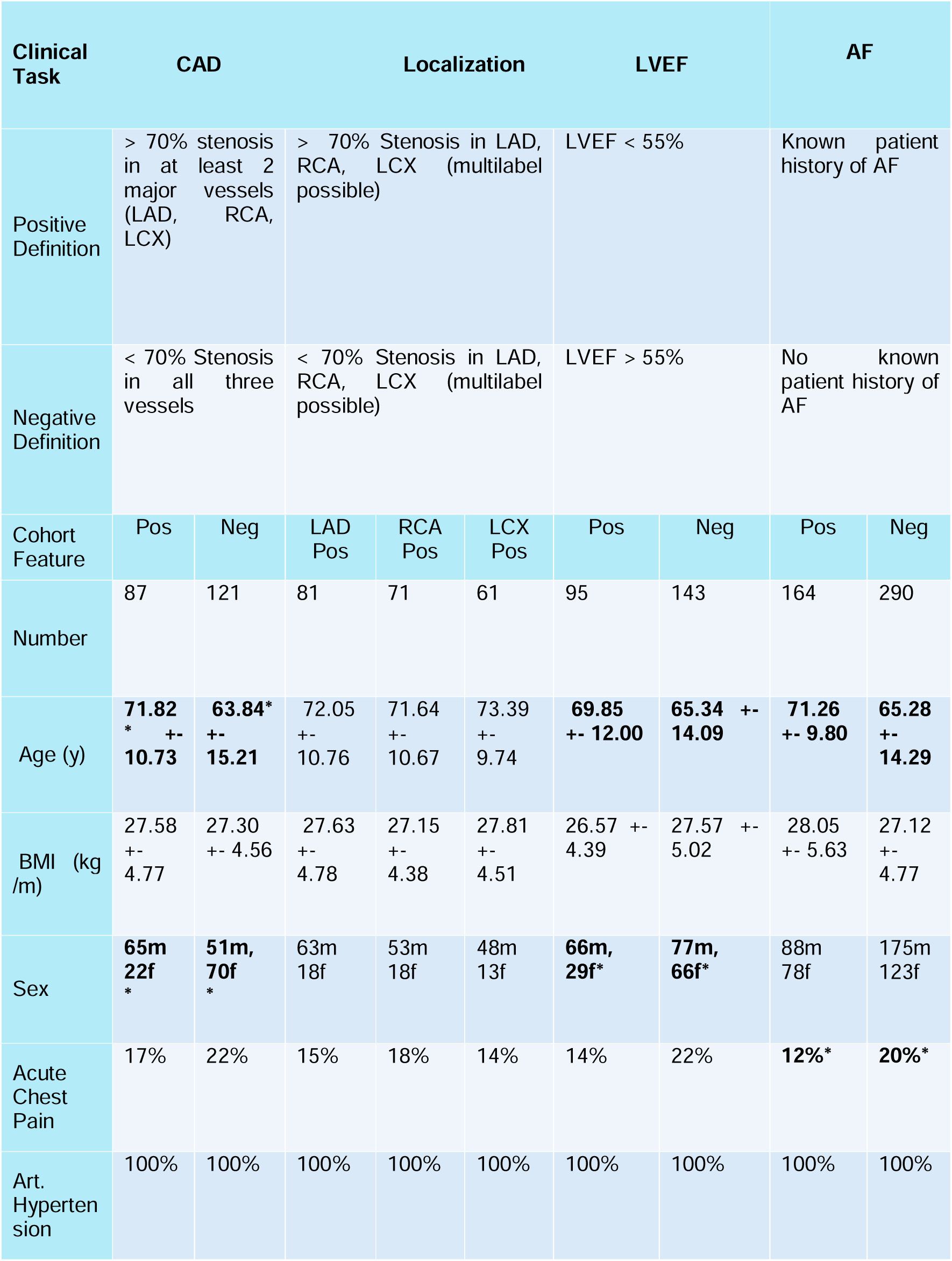
Clinical cohort description. Statistically significant (p < 0.05) distributional differences in bold (Mann-Whitney-U Test for continuous variables, χ^2^ for categorical variables).

### Clinical tasks

The pretrained MCG2Vec encoder demonstrated strong generalization across diverse clinical endpoints, each targeting a distinct aspect of cardiac pathophysiology. For the detection of significant CAD, the model achieved a mean AUC of 0.89 (95% CI: 0.82–0.93; Fig. 4a), indicating reliable discrimination between patients with angiographically confirmed multivessel stenosis and those without. Importantly, performance remained consistent across vessel-specific subanalyses (LAD, LCX, RCA; AUC = 0.84–0.89), supporting the encoder’s capacity to extract spatially localized ischemic signatures from non-invasive MCG (Fig. 4c-e). When fine-tuned to identify reduced LVEF (< 55%), the model reached an AUC of 0.81 (95% CI: 0.71–0.91; Fig. 3a), suggesting that the MCG signal encodes features reflective of global myocardial contractility. These findings align with the notion that magnetic field dynamics carry information about both depolarization and repolarization, potentially providing a functional correlate of ventricular performance. Finally, for AF classification, the model achieved an AUC of 0.77 (95% CI: 0.69–0.85; Fig. 4a), despite all recordings being acquired during sinus rhythm. This result indicates that MCG embeds subtle morphological or temporal markers of atrial remodeling and electrophysiological instability, even in the absence of active arrhythmia. Given the inherent label noise in clinical history–derived AF annotations, this level of performance highlights the robustness and potential clinical utility of the learned representations.

**Figure 4.**
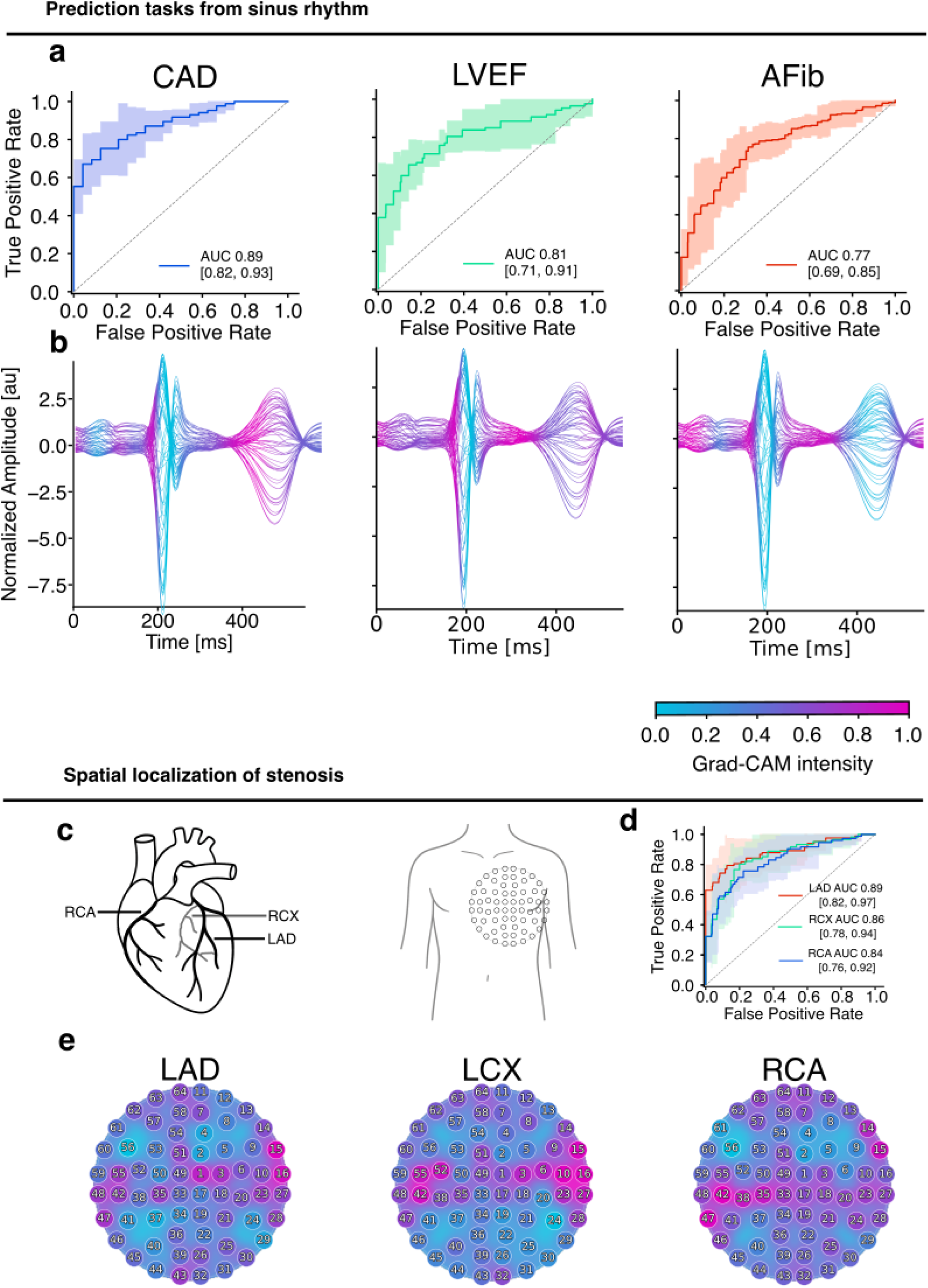
(**a**) Receiver operating characteristic (ROC) curves showing fine-tuned model performance for CAD, reduced LVEF, and AF classification, all derived from sinus rhythm recordings. Shaded areas indicate 95% confidence intervals. (**b**) Channel-wise averaged beats of the validation sets, color-coded by Grad-CAM intensity, highlighting morphology segments most relevant to model predictions. (**c**) Schematic of coronary arteries (LAD, LCX, RCA) and MCG sensor layout used for spatial localization analysis. (**d**) ROC curves for artery-specific stenosis localization tasks. (**e**) Spatial Grad-CAM activation maps across the 64-channel array for LAD, LCX, and RCA lesions, revealing distinct topographic signatures for each affected vessel.

### Explainability analysis

To validate the physiological basis of the model’s predictions, temporal and spatial activation maps were generated for the validation cohort. Representative beat-aligned importance profiles are shown in Figure 4b.

For the detection of CAD, the model’s attention was distributed across the cardiac cycle, with high-intensity activations observed during the beginning of the QRS complex and the ST-T wave segment. In contrast, for the prediction of reduced LVEF, salient features were temporally confined to the phase of ventricular depolarization, centering on the onset and peak of the QRS complex. The prediction of AF risk from sinus-rhythm recordings was associated with a distinct activation pattern focused on the terminal portion of the QRS complex (S-wave) and the P-wave.

Spatially, the projection of importance scores onto the 64-channel sensor array revealed distinct topographic signatures for vessel-specific ischemia. As shown in Figure 4e, the distribution of high-importance sensors varied according to the specific target vessel (LAD, LCX, or RCA), indicating that the model leveraged spatially localized magnetic field alterations to discriminate between ischemic territories.

## Discussion

This study demonstrates that a self-supervised deep learning framework, when applied to high-fidelity magnetocardiography, can extract rich diagnostic information from short, sinus-rhythm recordings. The proposed MCG2Vec encoder successfully generalized across distinct pathophysiological domains, detecting anatomical stenosis (CAD), functional impairment (reduced LVEF), and electrophysiological instability (AF risk). By combining the biophysical sensitivity of quantum sensors with the feature-learning capabilities of modern AI, our results suggest that MCG can bridge the long-standing diagnostic gap between accessible but limited surface electrocardiography and high-precision but resource-intensive tomographic imaging.

While recent advances in AI have enabled the detection of structural heart disease from standard 12-lead ECGs [24], [25], [26], these models remain constrained by the physics of the input signal. Electrical potentials are attenuated and smoothed as they traverse the thorax, acting as a low-pass filter that obscures fine-grained electrophysiological details [9]. Our findings underscore that MCG overcomes these limitations. Magnetic fields propagate through tissue with negligible distortion, preserving the complex spatiotemporal morphology of the cardiac field. Furthermore, theoretical models suggest that MCG is sensitive not only to transmembrane potentials but also to intracellular primary currents (vortex currents) associated with ionic flux [9], [11]. The model’s ability to detect ischemia and localize it to specific vascular territories (LAD, RCA, LCX) likely stems from this enhanced sensitivity to local repolarization heterogeneity, which remains subthreshold in surface potentials. Similarly, the accurate detection of AF risk from sinus rhythm confirms that the magnetic signal captures latent markers of atrial remodeling and fibrosis—a “magnetic phenotype” of the vulnerable substrate—before the onset of overt arrhythmia.

Recent systematic reviews have demonstrated broad applications of AI in cardiology—spanning risk prediction, imaging analysis, treatment optimisation, and clinical decision support [27], [28], [29]. Landmark studies using convolutional neural networks have even achieved or surpassed human-level accuracy in ECG-based arrhythmia detection [24] and automated 12-lead ECG diagnosis [25]. Most recently, Columbia’s EchoNext tool showed enhanced accuracy in detecting structural heart disease from standard ECGs [26]. Many prior AI studies in cardiology have been evaluated against clinically less relevant tasks, for example detecting atrial fibrillation from convenience ECGs in mixed hospital populations [30]. Such study designs risk overestimating performance, as models may exploit demographic or referral biases or detect general disease burden rather than disease-specific physiology. These limitations are not unique to atrial fibrillation; similar concerns have been raised across cardiovascular imaging, wearable sensing, and risk prediction studies, where unrepresentative cohorts and insufficient ground-truth verification limit generalizability [31] [32] [33]. Methodological reviews and reporting guidelines likewise emphasize that AI systems must be evaluated in realistic patient cohorts and clinically relevant diagnostic dilemmas to achieve meaningful impact [34]. For AI to meaningfully impact clinical pathways, evaluations must be performed in realistic patient cohorts where genuine diagnostic dilemmas exist.

A critical barrier to the clinical adoption of AI is the “black box” phenomenon. To address this, we employed a contrastive learning approach that forces the model to learn invariant morphological features rather than relying on patient-specific noise or channel artifacts. The Grad-CAM analysis provides validation of this approach: the model consistently focused on physiologically relevant segments of the cardiac cycle. For instance, attention was drawn to the ST-T segment for ischemia and the P-wave/S-wave boundaries for atrial fibrillation risk. These patterns align with established electrophysiological principles, providing a layer of interpretability that distinguishes this framework from purely statistical “black box” predictions.

The integration of AI-enabled MCG into the diagnostic workflow offers the potential to redefine non-invasive cardiology. Currently, the diagnostic pathway for chest pain or heart failure often involves a cascade of tests, radiation exposure, and contrast agents [4]. An AI-enhanced MCG assessment could serve as a rapid, contactless, and radiation-free “gatekeeper” at the point of care. In patients with intermediate pre-test probability, a negative MCG screening could reduce unnecessary angiograms, while a positive result could expedite downstream therapy.

Beyond triage, this technology paves the way for a shift from reactive to preventive cardiology. Because MCG requires no contact or preparation, it is uniquely improving longitudinal monitoring. Once an initial diagnostic baseline is established, follow-up assessments could rely on serial MCG recordings to detect subtle electrophysiological deviations—essentially a “digital twin” approach. This would allow for the monitoring of disease progression or therapeutic response in heart failure and CAD without the logistical and economic burden of repeated echocardiography or nuclear imaging.

### Limitations

Our study has several limitations. First, the dataset, while large for an MCG study, is retrospective and single-center. Second, atrial fibrillation status was determined based on clinical history. While this reflects real-world phenotyping, it lacks the precision of continuous rhythm monitoring, and it is possible that some “control” patients had undiagnosed paroxysmal AF. Third, approximately 75% of the recordings were acquired with a hardware bandpass filter. While our contrastive pre-training proved robust to these variations, future studies using unfiltered “DC” MCG data might yield even higher diagnostic accuracy by preserving these frequency components of the magnetic field. Finally, the reference standard for ischemia was anatomical stenosis; future work should correlate MCG findings with functional ischemia (FFR) and microvascular parameters to fully elucidate the structure-function relationship.

### Conclusion

We introduce an AI-enabled magnetocardiography framework that achieves high diagnostic accuracy for coronary, ventricular, and rhythmic disorders using a single, non-invasive resting scan. by unlocking the high-dimensional information within the cardiac magnetic field, this approach overcomes the biophysical limitations of the ECG and offers a scalable, radiation-free adjunct to precision cardiac diagnostics.

Beyond the methodological and diagnostic implications, the integration of AI and MCG has the potential to fundamentally transform the clinical workflow in cardiology. In a typical diagnostic pathway, patients presenting with chest pain or cardiovascular risk factors undergo a cascade of investigations like ECG, TTE, laboratory tests, and, if indicated, stress testing or advanced imaging. Introducing an AI-enabled MCG at the initial assessment provides a rapid, contactless, and radiation-free procedure that complements conventional findings while adding precise electrophysiological insight into cardiac function and ischemic risk.

Once an initial diagnostic baseline is established, follow-up assessments could rely primarily on serial MCG recordings, reducing the need for repeated imaging, laboratory testing, or functional stress procedures. Such an approach would not only streamline patient management but also decrease diagnostic redundancy, lower costs, and relieve pressure on healthcare personnel. Importantly, AI-driven longitudinal comparison of MCG signatures could allow the early detection of electrophysiological deterioration, enabling timely intervention and shifting the diagnostic paradigm from reactive to preventive cardiology.

The convergence of quantum sensing, deep learning, and clinical cardiology may thus mark a transition from static testing toward dynamic, personalized disease surveillance.

Taken together, AI-enabled MCG may open a new diagnostic frontier in cardiology by uniting electrophysiology, imaging, and machine learning into a unified, non-invasive diagnostic continuum that aligns with the goals of precision and preventive medicine.

## Data Availability

All data in the present study are available upon reasonable request to the authors

